# Breaking down the costs for breast cancer: Insights from Sweden’s National Quality Register

**DOI:** 10.1101/2025.06.24.25330188

**Authors:** Xiaoyang Du, Leo Gkekos, Balram Rai, Anna L.V. Johansson, Irma Fredriksson, Mattias Rantalainen, Emelie Heintz, Shuang Hao, Mark Clements

## Abstract

**Background:** With emerging new technologies for diagnostics and treatment for breast cancer, there is a demand for updated breast cancer costs based on current clinical practice. The objectives of the current study were to (a) estimate recent societal costs of breast cancer in Sweden and (b) provide population-based patient-level cost estimates for health economic evaluations.

**Methods:** This prevalence-based cost-of-illness study was based on 2019 data linking multiple Swedish national registers. The analysis employed a societal perspective considering direct health care, informal care, and productivity losses. Total costs were estimated using a bottom-up micro-costing approach. Direct costs were also estimated by subgroups, including age group, molecular subtype, breast cancer stage, and disease state defined by metastatic stage.

**Findings:** 82,960 breast cancer patients diagnosed since 2008 were alive by the end of 2019. The annual societal cost of breast cancer in Sweden was 526 million euro, where the direct health care, informal care, and productivity losses accounted for 54%, 7%, and 40%, respectively. Costs of direct health care, including inpatient/outpatient care and prescribed drugs, varied by subgroups, where younger age, higher stage, and more adverse subtypes were associated with higher costs per patient-year. Patients with a diagnosis of de novo metastatic cancer incurred the highest cost per patient-year.

**Interpretation:** Breast cancer represents a large economic burden in Sweden. The mean cost estimates per patient-year are informative to future health economic evaluations for breast cancer screening and treatment.

## Introduction

Breast cancer is the most diagnosed cancer and comprised 13.8% of all cancer cases among European Union (EU) countries.^1^ In Sweden, breast cancer is the most common cancer for women, with a 10-year prevalence of 63,017 cases in 2022.^2^ Incidence of breast cancer in Sweden increased between the 1960s and 2000s and has been stable over the past 15 years, while mortality has declined.^2^ A potential major contributor for the reduction in mortality is the early detection through mammography screening.

National breast cancer screening in Sweden was introduced in the 1980s. Since 2016, mammographic screening has been free of charge for women aged 40 to 74 years. Several Swedish studies have assessed the effectiveness of breast cancer screening at the regional and national level and found that increased screening was associated with reductions in the incidence of advanced cancer and of breast cancer mortality.^3,4^ For metastatic disease, patient survival may be prolonged due to the introduction of advanced treatment strategies using innovative targeted drugs, although these drugs may be expensive and could increase health care expenditure.^5,6^

The economic burden of a disease can be assessed through a cost-of-illness (COI) study.^7^ Two studies have investigated the costs of breast cancer in Sweden from a societal perspective including both direct and indirect costs. Lidgren and colleagues estimated the costs of prevalent breast cancer cases diagnosed from 1958 to the end of 2002.^8^ By using a top-down method, the estimated total cost was 3 billion Swedish Kronor (SEK), with the majority of the costs attributed to productivity losses. Luengo-Fernandez and colleagues estimated the cancer costs for 2009 using aggregate data at the European level.^9^ The breast cancer costs were estimated at approximately 280 million euro for Sweden. With emerging new technologies for diagnostics and treatment, these data are less informative. There is a demand for updated results based on current clinical practice. In addition, previous Swedish studies on breast cancer costs by different subgroups, such as by breast cancer subtype and by disease state, were conducted on small sample sizes in single clinics, and the findings may not be generalisable to a broader population.^10,11^ By using the latest available linked register data, our study aimed (a) to update the total cost of breast cancer in Sweden to reflect the most recent management of breast cancer, and (b) to estimate the average costs per patient by various subgroups, to inform future health economic evaluations of screening and interventions targeting breast cancer.

## Method

This was a register-based, prevalence-based COI study for breast cancer in Sweden. A societal perspective was applied, where both direct costs and indirect costs were included. Individual-level resource health-care utilisation in 2019 was extracted from the Breast Cancer Database Sweden (BCBaSe) 3.0.^12^ For care that was not included in BCBaSe 3.0, resource use was estimated using information from governmental reports, existing literature and a European-level survey for informal care (Figure 1). Unit costs from year 2023 were applied if available in price lists. Otherwise, unit costs for other years were adjusted to the price level of 2023 using the consumer price index. Total costs were calculated using a bottom-up method through the multiplication of the quantities of resources by the unit costs and converted to euro (€1 = 11.48 SEK, 2023 rate).^13^ Direct costs for inpatient/outpatient care and prescribed drugs were estimated for different subgroups, including age group, breast cancer subtype, disease stage at diagnosis defined by the TNM classification, and disease state defined by metastasis status. Analyses were performed using R (version 4.2.3, R Foundation for Statistical Computing, Vienna, Austria) and Stata/BE 17.0 (Corp, College Station, Texas, USA).

**Figure 1.**
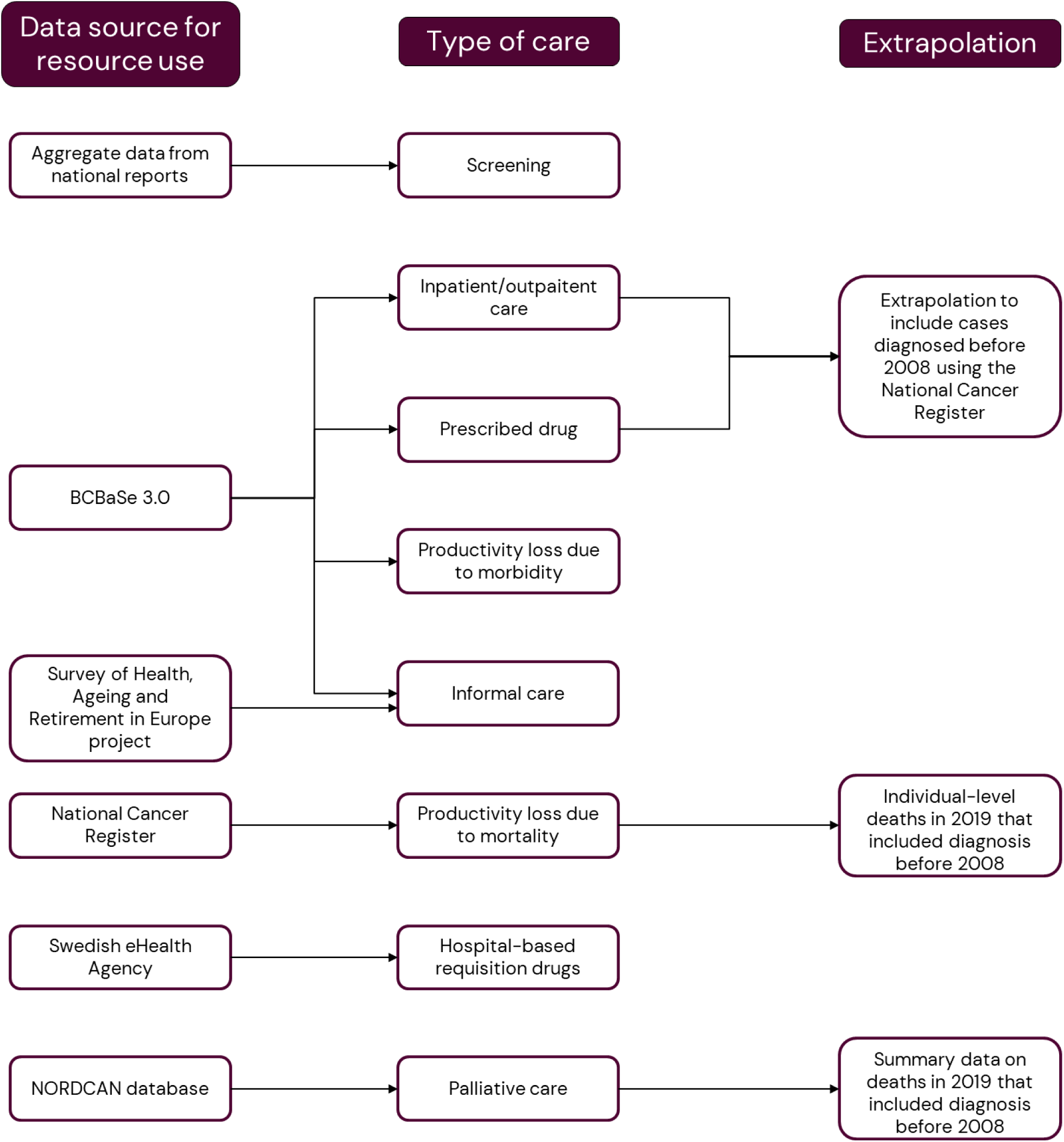
Data source for resource use in each type of care.

### Study population base

The study included (a) women invited for a mammography screening in 2019 and (b) individuals diagnosed with invasive or non-invasive breast cancer (C50 & D05, The International Classification of Diseases, 10th version, ICD-10) prior to 31 December 2019 in Sweden. The diagnosed individuals were retrieved through BCBaSe 3.0, which is a research database that has linked ten registers and has national coverage for breast cancer cases diagnosed from 2008 in Sweden. The linked registers include the Swedish National Breast Cancer Quality Register, the Total Population Register, the National Cause of Death Register, the National Cancer Register, the National Patient Register, the National Prescribed Drug Register, the Medical Birth Register, the Multi-Generation Register, the Longitudinal Integrated Database for Health Insurance and Labor Market Studies, and the Micro Data for Analysis of Social insurance (MiDAS database). To extrapolate the costs to all prevalent cases in Sweden, we used cancer registrations from the NORDCAN database and the National Cancer Register to complement the breast cancer cases diagnosed before 2008 (Figure 1).^2^

### Resources utilisation and costs

#### Screening

Women aged between 40 and 74 years in Sweden are invited for a mammography screening every 18-24 month. Using aggregate-level data from the National Board of Health and Welfare, 78% of invited women participated in mammography screening in 2019-2020.^14^ We assumed that half of the screening population were screened in 2019. According to the national mammography screening guidelines, 3% of women who participated in the screening were recalled for further investigation.^15,16^ After clinical mammography, breast and axillary ultrasound, 33% would have an ultrasound-guided core biopsy and/or fine needle aspiration. The price of each procedure was extracted from a report by the Institute for Health Economics.^15^ The total costs were calculated as a product of the number of participants for each procedure multiplied by the unit cost.

#### Inpatient and outpatient care

Inpatient and outpatient care episodes were identified using the Nordic Diagnostic Related Groups (DRG), given a major diagnosis category (MDC) code of 23 (other and unspecified health problems), 30 (breast diseases) or 40 (overall problems in outpatient care), with a diagnosis of C50 or D50 in BCBaSe 3.0 (Supplementary Table 3). The unit costs for each DRG code were extracted from the Swedish Association of Local Authorities and Regions.^17^ The total costs were calculated by multiplying the number of DRGs by their respective unit costs. To estimate the cost of diagnostic procedures for the patients who were clinically diagnosed, we included relevant DRG codes prior to a diagnosis of breast cancer (for details of the method, see Supplementary Appendix A5).

#### Pharmaceuticals

We identified 36 substances with indications for breast cancer that had been approved for the Swedish market by 2019 (Supplementary Table 5). The quantities of drug prescriptions in 2019 were extracted from BCBaSe 3.0. Unit costs of drugs by brand and package were identified through The Dental and Pharmaceutical Benefits Agency database or from pharmacy websites (for details, see Supplementary Appendix A6). The costs for the prescribed drugs were calculated from the number of prescriptions multiplied by the unit costs for each prescription. Hospital-based requisition drugs were not included in the BCBaSe 3.0. Aggregated costs for requisition drugs in 2019 were retrieved from the Swedish eHealth Agency and adjusted to 2023 values. Supportive drugs for chemotherapy and drugs for adverse effects of breast cancer medications and for skeletal metastasis were not included in this study.

#### Palliative care

We identified breast cancer deaths in 2019 from the NORDCAN database and assumed that all patients who died had received palliative care. We assumed that 10.5% of these patients resided in nursing homes based on a Swedish register study.^18^ Given a recent study from the Stockholm region, we used the mean costs for palliative care per patient with cancer for nursing-home residents and for non-nursing-home residents.^19^ Total costs for palliative care were calculated using the number of death cases multiplied by the unit cost per patient.

#### Informal care

Resource use of informal care was estimated using data from the Survey of Health, Ageing and Retirement in Europe (SHARE) project.^20^ We extracted data from WAVE8, which was the last survey preceding the COVID-19 pandemic. We used logistic regression to model the probability that a patient received informal care from a working-age (aged less than 65 years) caregiver for (a) patients who were severely limited in daily activities and (b) patients with terminal illness. For patients who received informal care from a working-age caregiver, the number of hours was modelled using linear regression. Informal care was valued at 28.5 euro per hour based on the average salary (including social security fees) in Sweden in 2023.^21^ The total costs for informal care were calculated by multiplying the predicted number of hours and the cost per hour. Further details are provided in Supplementary Appendix A7.

#### Productivity losses

Productivity losses due to morbidity and pre-mature mortality were estimated using the human capital method. For productivity losses due to morbidity, we identified the patients who had received sick leave benefits and disability pension due to a breast cancer diagnosis as a main cause (C50 & D05) from the Swedish Social Insurance Agency in BCBaSe 3.0. In Sweden, a short-term sick leave up to 14 days is paid by the employer, while a longer-term sick leave is covered by the Swedish Social Insurance Agency. In addition, temporary and permanent disability pensions for individuals whose work capacity was considered impaired were recorded. We extracted the number of net days that a patient had been receiving sick leave benefits and disability pensions from BCBaSe 3.0. Sick leaves within 14 days were not recorded and thus not included in our calculations. However, patients who had received sick leave payments from the Swedish Social Insurance Agency were assumed to have the 14-day sick leave beforehand. We valued the lost productivity until 65 years old (the general retirement age in Sweden) and applied a mean cost of 228 euro per day, which was calculated based on the average gross earning including social security fee in 2023 in Sweden.^21^ The total cost was calculated using the number of net days multiplying by the unit cost per day.

For analysis of productivity losses due to premature death, deaths due to breast cancer in 2019 were identified in the National Cancer Register. the accumulated losses of years up to age 65 years were calculated using the background mortality from the Swedish general population together with a 3% discount rate for future productivity losses. The details of the method are described in Hao et al.^22^

### Subgroup analysis

#### Breast cancer subtypes

Breast cancer subtypes were defined based on estrogen receptor (ER) status, progesterone receptor (PR) status, human epidermal growth factor receptor 2 (HER2) status, and Nottingham histologic grade (NHG). Expression ≥ 10% was defined as positive for both ER status and PR status in accordance with Swedish treatment guidelines (Supplementary Table 1). For patients with more than one diagnosis of breast cancer, we restricted to the subtype of the first diagnosis. Costs of inpatient/outpatient care and prescribed drugs were reported by subtype.

#### Disease states

Disease states were defined as (a) primary diagnosis without distant metastases (M0), (b) primary diagnosis with de novo metastasis (M1), and (c) distant recurrence after primary diagnosis without metastasis. Multiple imputation was performed for missing M stage at diagnosis and for missing date of distant recurrence for breast cancer death cases. For details on the imputation process, see Supplementary Appendix A3. Costs of inpatient/outpatient care and prescribed drugs were reported by disease state.

#### Breast cancer stages

Breast cancer stages were defined using the TNM classification 8^th^ edition (Supplementary Table 2).^23^ For patients with more than one diagnosis of breast cancer, we restricted to the TNM status of the first diagnosis. The analysis was performed using the imputed datasets for M stage. Costs of inpatient/outpatient care and prescribed drugs were reported by stage.

For the analysis of mean costs per patient-year by different subgroups, we undertook a period analysis for 2019 with possible left truncation and right censoring for time since diagnosis, and for disease state, additionally split time since distant recurrence (for illustration of the method, see Supplementary Appendix A4). The average costs were calculated as the accumulated costs since diagnosis (and since distant recurrence for disease state) divided by the accumulated person-years in 2019. For analysis of the disease states, we calculated the confidence intervals for the mean costs per patient-year, which were based on the normal variance of the observed costs together with the binomial variance of the number of patients with costs.

### Extrapolation to the prevalent cases in Sweden

We extrapolated the costs of inpatient/outpatient care and prescribed drugs to include prevalent breast cancer cases diagnosed in Sweden before 2008. We performed the same period analysis for time since diagnosis using data from the Swedish Cancer Register and calculated the accumulated patient-years by age group and by year since diagnosis to 2019. Average costs by patient-year were based on BCBaSe 3.0 data. The total costs were summed by multiplying the accumulated person-years from the Swedish Cancer Register with average costs by patient-year.

### Sensitivity analysis

One-way sensitivity analyses were performed in the following scenarios: (a) for inpatient/outpatient care, we used resource use data in the KPP (Kostnad per patient, cost per patient) system from the Swedish Association of Local Authorities and Regions; (b) we used the proxy good method to estimate costs for informal care, where it was assumed that the care provided by informal caregivers is valued at the market price of formal caregivers regardless of their working age. An hourly cost of 20.7 euro was applied based on the average gross earning including social security fee of a personal care worker in health services in 2023.^21^

## Results

### Study population

According to the data in BCBaSe 3.0, there were 82,960 patients living with a diagnosis of breast cancer in Sweden by the end of 2019 (Table 1). Among 8,734 patients diagnosed in 2019, 4,291(49%) were detected through screening. Of the 952 deaths due to breast cancer that occurred in 2019, more than half were in patients aged ≥70 years.

**Table 1.** Description of the BCBaSe 3.0 study population (Sweden 2019)

| **Age group** | **Prevalent cases** | **Incident cases** | **All-cause deaths** | **Breast cancer deaths** |
| --- | --- | --- | --- | --- |
|  | **N (%)** | **N (%)** | **N (%)** | **N (%)** |
| <40 | 3,308 (4) | 335 (4) | 20 (1) | 18 (2) |
| 40-49 | 13,144 (16) | 1,215 (14) | 83 (3) | 72 (8) |
| 50-59 | 17,944 (22) | 1,618 (19) | 177 (7) | 125 (13) |
| 60-69 | 26,063 (31) | 2,258 (26) | 335 (13) | 173 (18) |
| ≥70 | 22,501 (27) | 3,308 (38) | 1,985 (76) | 564 (59) |
| Total | 82,960 (100) | 8,734 (100) | 2,600 (100) | 952 (100) |

### Screening

Given the assumption that 78% of invited women participated in breast cancer screening, we estimated 705,913 women to have undergone screening mammography during 2019, of whom 21,177 were estimated to be selected for a clinical work-up with 7,059 women to have undergone a biopsy. The estimated total costs of breast cancer screening were 63 million euro (Table 2).

**Table 2.** Costs of breast cancer by type of care in Sweden (2023 price, euro)

| Type of care | Cost (€) | Percentage of total cost (%) |
| --- | --- | --- |
| **Direct cost** | **281,905,564** | **53.63%** |
| Screening | 62,663,694 | 11.92% |
| Inpatient care | 39,259,457 | 7.47% |
| Outpatient care | 72,841,119 | 13.86% |
| Pharmaceuticals | 66,747,738 | 12.70% |
| Palliative care | 40,393,556 | 7.68% |
| **Informal care** | **34,251,936** | **6.52%** |
| Severely limited in daily activities | 29,763,136 | 5.66% |
| Terminal illness | 4,488,800 | 0.85% |
| **Productivity loss** | **209,484,902** | **39.85%** |
| Morbidity- Sick leave | 184,679,943 | 35.13% |
| Morbidity- Disability pension | 8,103,690 | 1.54% |
| Premature mortality | 16,701,269 | 3.18% |
| **Total cost** | **525,642,401** | **100%** |

### Inpatient and outpatient care

In BCBaSe 3.0, there were 4,618 patients admitted to inpatient care for breast cancer in 2019 for 5,349 episodes. The most frequent DRG code for inpatient care was total mastectomy for malignant tumor, accounting for 36% of the total cost of inpatient care for breast cancer (Supplementary Table 7). There were 16,762 patients who attended outpatient care with 91,456 episodes. The most frequent DRG code for outpatient care was “doctor visits for malignant breast diseases”, followed by “less resource-intensive radiotherapy” (Supplementary Table 7). The diagnostic and care procedures prior to a diagnosis for clinically detected cases contributed 0.2 million euro in 2019. After extrapolation to all prevalent cases in Sweden, the total costs of inpatient care and outpatient care were estimated to be 39 million euro and 73 million euro respectively (Table 2).

### Pharmaceuticals

Based on BCBaSe 3.0, 36,218 patients had 26 substances prescribed for breast cancer treatment in 2019. The most prescribed substance was tamoxifen, followed by letrozole and anastrozole (Supplementary Table 7). Palbociclib had the highest cost (5.7 million euro), followed by letrozole (2.3 million euro) and goserelin (1.3 million euro). By drug class, CDK4/6 Inhibitors had the highest cost at 6.4 million euro, accounting for 46% of the total prescribed drugs costs in BCBaSe 3.0 (Supplementary Table 8). After extrapolation to all prevalent cases in Sweden, the estimated cost of prescribed pharmaceuticals was 21 million euro. For hospital requisition drugs, 20 and 35 substances were extracted from the Swedish eHealth Agency for outpatient and inpatient care, respectively. We included the drugs with only indication for breast cancer and the estimated cost was 46 million euro.

### Palliative care

According to the NORDCAN database, there were 1,365 breast cancer deaths in 2019. Based on the assumption that all patients who died in 2019 had received palliative care and 144 of them were nursing-home residents, the total cost of palliative care was estimated to be 40 million euro (Table 2).

### Informal care

We included 74,352 patients that were diagnosed before 2019 and were alive in 2019 from BCBaSe 3.0 as the study base for receiving informal care due to their limitation in daily activities. A total number of 1,044,321 hours of care provided from inside and outside the household were estimated, yielding a total cost of 30 million euro (Table 2 and Supplementary Table 7). We included 952 patients in BCBaSe 3.0 who died in 2019 and assumed that they received informal care due to terminal illness. The total cost for 157,502 hours of informal care for terminal illness was estimated to be 4.5 million euro (Table 2 and Supplementary Table 7).

### Productivity losses

The cost of productivity losses due to morbidity were estimated to be 199 million euro (Table 2). In BCBaSe 3.0, 5,690 patients had a period of sick leave with a total number of 832,055 net days in 2019, which contributed to 96% of the productivity losses due to morbidities (Supplementary Table 7). There were 145 patients who received disability pension from the Swedish Social Insurance Agency, with the cost of productivity losses due to morbidities being 8 million euro.

According to the National Cancer Register, there were 352 patients aged < 65 years who died from breast cancer in 2019. Productivity losses due to premature mortality were estimated to a cost of 17 million euro (Table 2).

### Costs by subgroup

The highest mean costs per patient-year in the first year following a primary diagnosis were seen for patients aged < 40 years (Table 3). The mean cost decreased with increasing age. Patients with HER2+ disease had the highest mean costs in the first year after a primary diagnosis, followed by those with luminal HER2+ cancer and those with triple negative cancer (Table 3). In the first year since a diagnosis, patients at stage III had the highest mean cost (14,974 euro per patient-year). However, since the second year, patients at stage IV had the highest cost at 6,078 euro per patient-year (Table 3), while the costs dropped to below 1100 euro per patient-year for patients at stage I-III (Table 3). Patients diagnosed with de novo metastatic breast cancer had the highest mean cost in the first year as well as in the subsequent years after diagnosis (Table 3 and Figure 1).

**Table 3.** Direct Costs combining inpatient/outpatient care and prescribed drugs, by subgroups (2023 price, euro)

| Subgroup | First year of follow-up |  |  | Second and subsequent years of follow-up |  |  | Total years of follow-up |  |  |
| --- | --- | --- | --- | --- | --- | --- | --- | --- | --- |
|  | Total cost (€) | Total patient-years | Cost/patient-year (€) | Total cost (€) | Total patient-years | Cost/patient-year (€) | Total cost (€) | Total patient-years | Cost/patient-year (€) |
| **Breast cancer subtype** |  |  |  |  |  |  |  |  |  |
| Luminal A like | 33,375,236 | 3,462 | 9,642 | 8,690,843 | 32,331 | 269 | 42,066,079 | 35,793 | 1,175 |
| Luminal B like | 13,559,611 | 1,200 | 11,300 | 4,181,243 | 10,202 | 410 | 17,740,854 | 11,402 | 1,556 |
| Luminal HER2+ | 9,258,767 | 680 | 13,616 | 2,826,176 | 5,384 | 525 | 12,084,943 | 6,064 | 1,993 |
| HER2+ | 4,266,755 | 301 | 14,180 | 887,248 | 2,419 | 367 | 5,154,003 | 2,720 | 1,895 |
| Triple negative | 8,720,981 | 671 | 12,992 | 1,541,160 | 4,559 | 338 | 10,262,141 | 5,230 | 1,962 |
| **Breast cancer stage** |  |  |  |  |  |  |  |  |  |
| Stage 0 | 4,277,268 | 561 | 7,624 | 978,539 | 5,014 | 195 | 5,194,317 | 5,575 | 941 |
| Stage I | 31,967,902 | 3,506 | 9,119 | 8,033,018 | 33,365 | 241 | 39,075,689 | 36,870 | 1,085 |
| Stage II | 38,523,974 | 3,255 | 11,837 | 11,456,346 | 24,217 | 473 | 47,674,412 | 27,472 | 1,819 |
| Stage III | 10,034,559 | 670 | 14,974 | 5,722,829 | 5,449 | 1,050 | 13,904,666 | 6,119 | 2,575 |
| Stage IV | 2,605,054 | 191 | 13,620 | 3,420,449 | 563 | 6,078 | 4,556,952 | 754 | 7,991 |
| **Age group** |  |  |  |  |  |  |  |  |  |
| <40 | 4,692,820 | 283 | 16,555 | 1,246,168 | 999 | 1,248 | 5,938,988 | 1,282 | 4,632 |
| 40-49 | 14,808,300 | 1,102 | 13,441 | 3,794,796 | 5,342 | 710 | 18,603,096 | 6,444 | 2,887 |
| 50-59 | 18,524,942 | 1,603 | 11,556 | 6,191,493 | 13,193 | 469 | 24,716,435 | 14,796 | 1,670 |
| 60-69 | 22,230,516 | 2,125 | 10,460 | 6,963,166 | 17,390 | 400 | 29,193,682 | 19,515 | 1,496 |
| ≥70 | 28,614,990 | 3,231 | 8,856 | 11,691,711 | 32,494 | 360 | 40,306,701 | 35,725 | 1,128 |
| **Disease state** |  |  |  |  |  |  |  |  |  |
| Primary diagnosis M0 | 85,806,509 | 8,122 | 10,565 | 19,250,593 | 67,802 | 284 | 105,057,103 | 75,924 | 1,384 |
| De novo metastatic M1 | 2,605,054 | 191 | 13,620 | 3,420,449 | 563 | 6,078 | 6,025,503 | 754 | 7,991 |
| Distant recurrence | 3,566,038 | 363 | 9,820 | 4,110,262 | 721 | 5,697 | 7,676,298 | 1,085 | 7,078 |

### Sensitivity analysis

The change on the total costs in the sensitivity analyses ranged from −1.2% to 1.2% (Supplementary Figure 2). Using resources data from the KPP system led to minor decrease in the total costs of inpatient and outpatient care. Cost of informal care estimated by the proxy good method increased by 1.2% compared to base case.

## Discussion

Our study quantified the economic burden due to breast cancer using individual-level data in 2019 retrieved from BCBaSe 3.0 and extrapolated to all prevalent cases in Sweden. In total, the estimated annual social cost of breast cancer in Sweden was 526 million euro (Table 2). The estimated cost (excluding screening) per patient was 5581 euro and the estimated cost per capita was 51 euro in Sweden.

The estimated total cost is higher than presented in the previous economic burden studies of breast cancer in Sweden. In addition to inflation and price changes over time, the difference can be explained from several aspects. Firstly, compared to the Swedish study in 2002, our study also included palliative and informal care. This can partly explain the difference in the proportion of each cost component. In the previous Swedish study using the same human capital method as ours, indirect costs constituted 70% of the total cost, whereas in the present study the indirect costs accounted for approximately 40%.^8^ Another potential contributor to the proportional decline of productivity losses is the cost reduction due to premature mortality. Premature mortality accounted for 37% of the total cost in the 2002 study, whereas it explained only 3% in the present study. Improved treatment is expected to increase survival and life-expectancy.^24^ Our proportion of productivity losses was close to the European-level study (41%), although their calculation set 79 years as the general retirement age.^9^ Secondly, we used a bottom-up costing approach on individual-level data from the linked registers. As a strength of this study, this bottom-up method provides more precise estimates and allows a break-down of costs by subgroup compared to the top-down method or using aggregated data as per the previous studies.^7,25^ For example, we extracted individual-level DRG episodes from BCBaSe 3.0 rather than applying the aggregate cost in the KPP system. Compared to KPP, our study applied broader patient inclusion criteria. For our study, the DRG episodes were identified using an MDC code with any diagnosis of breast cancer rather than restricting to a primary diagnosis of breast cancer, e.g., patients with a primary diagnosis of metastatic cancer originating from breast were also included in the analysis.

Another strength of our study is the reporting of patient-level cost estimates by subgroup, including age group, breast cancer subtype, disease stage at diagnosis, and disease state, for direct health care costs comprising inpatient/outpatient care and prescribed drugs, which are informative for future economic evaluations. We found that younger patients were associated with higher costs. Patients aged < 40 years at diagnosis had a higher proportion of HER2+ and triple negative breast cancers subtypes compared to older groups (Supplementary Tables 6), requiring multimodal and more intensive treatments; thus, a higher health care expenditure is expected.

Costs by subtype have previously been investigated in the Swedish clinical setting; among 53 patients who died in year 2005-2006, patients with HER2+ disease had the highest mean cost,^10^ which is consistent with our findings. The mean cost differences among subtypes in our study were small. In contrast, studies in Canada and Portugal have shown a surge in the average costs among patients with HER2+ cancer.^26,27^ The cause of this discrepancy is unclear but may be driven by differences in the inclusion of type of care and assumed treatment duration. For example, compared to our study, the Canadian study included home care and palliative care, and results were reported per case given stage and treatment durations instead of per patient-year. Additionally, in the Canadian study, the largest rise in the cost for patients with HER2+ cancer was only for stage IV and due to cost changes in systemic therapy, while the cost differences between stages in other subtypes were smaller.^27^ It should be noted that, our study had a significant number of patients with unknown subtype (20%) that were excluded from the analysis. The reporting of HER2-testing was not completed until 2015. However, one could argue that the missingness may not be at random since HER2 testing was less often in older women diagnosed with cytology and primary endocrine therapy. Imputation on missing subtype should be further studied. Costs by stage have been widely investigated in several healthcare settings, showing that higher stage is associated with higher costs.^28,29^ To the best of our knowledge, our study is the first to report stage-specific breast cancer costs in the Swedish population. In our study, patients at stage III had the highest initial cost per patient-year following a primary diagnosis. However, at stage IV, the cost was significantly higher than the costs at the lower stages in the subsequent years, which was contributed mostly by prescribed drugs (Supplementary Table 9).

Lidgren and colleagues investigated disease state-specific costs for breast cancer in a single clinic in Sweden, including both direct costs and indirect costs.^11^ Based on more recent data, our study showed a similar pattern as Lidgren and colleagues, where costs dropped from the first to the second year in a given disease state. The highest costs were observed in the first year following a diagnosis of de novo metastatic breast cancer. For patients with non-metastatic breast cancer, most health care events and higher total costs emanated from outpatient care. There were marked differences in prescribed drug costs between different disease states, most likely due to the use of expensive targeted drugs among patients in the metastatic setting (Supplementary Table 9). For disease states defined by a distant recurrence or de novo metastatic cancer, the number of patients who had had episodes in hospitals or prescribed drugs decreased over time, leading to imprecise estimates. For example, a drastic increase in the prescribed drug costs was observed in the last few years of follow-up for very few patients with de novo metastatic cancer. Consequently, we truncated the follow-up for those disease states to 6 years (Figure 2).

**Figure 2.**
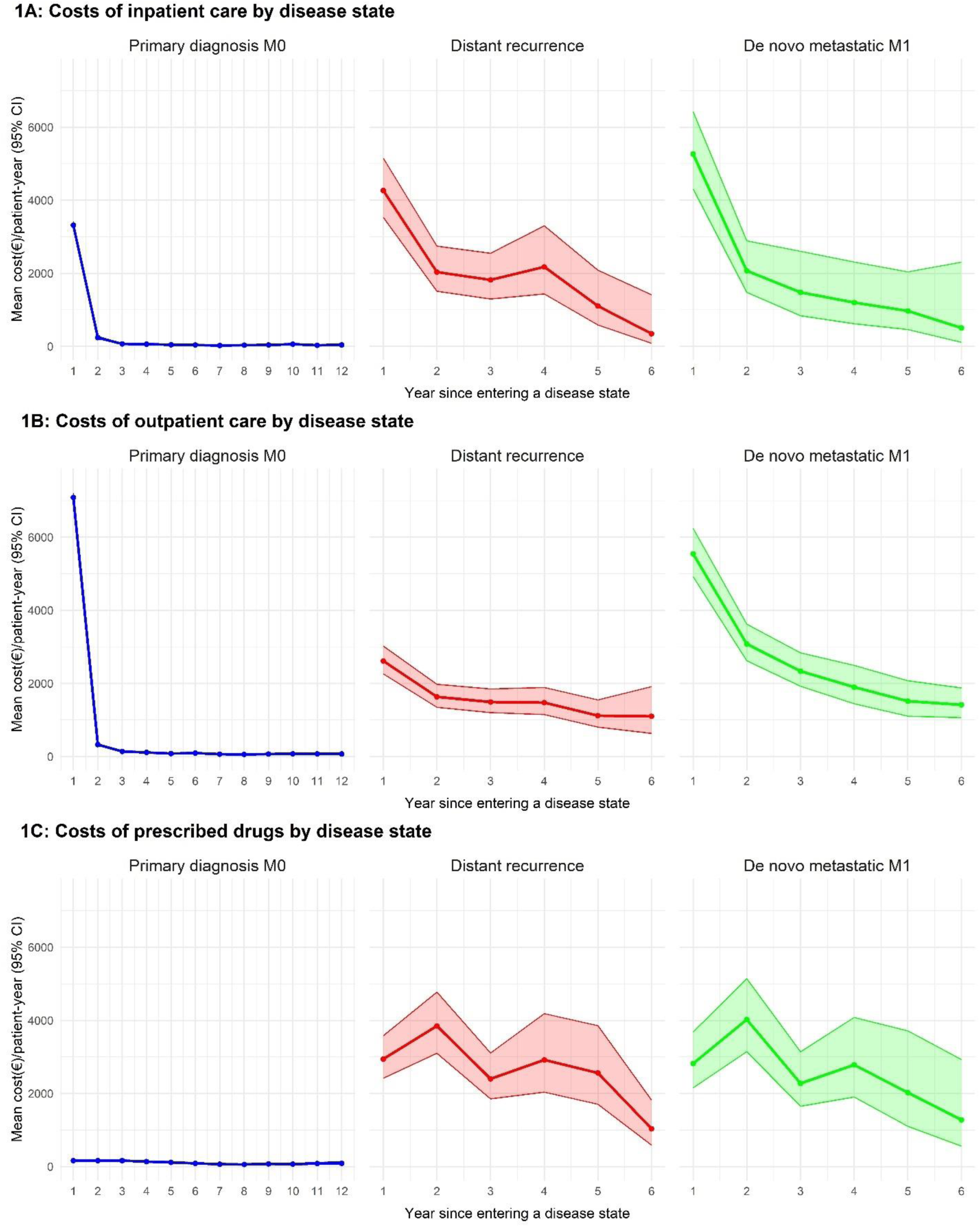
Direct cost (€)/patient-year by disease state. CI: Confidence interval.

Some limitations should be noted in our study. Firstly, resource use for screening, informal care and palliative care were estimated based on existing research. Estimates of screening attendance rates were made based on a national report. Among those who would be recalled for clinical mammography, around 20% would be diagnosed with breast cancer.^4,16^ The estimated number of diagnosed cases was close to the number of screening-detected cancer cases observed in BCBaSe 3.0 (4,236 vs. 4,291), which indicates the robustness of our assumptions. An empirical analysis was performed to estimate costs of diagnostic procedures for clinical diagnosed cases. Procedure costs for benign cases (with no diagnosis of C50 or D05) and costs for molecular diagnostics through gene expression assays were not included in the analysis. Consequently, the diagnostic costs will be underestimated. Informal care in our study cost less than informal care in the European study using a similar approach. In WAVE8 of the SHARE survey, the actual number of hours of care per day were not reported for those patients severely limited in daily activities. To address this issue, we used estimates from a previous study based on WAVE2. In addition, our calculations for informal care only included patients from BCBaSe 3.0, not including primary breast cancer diagnoses before 2008. Moreover, we did not link the Palliative Care Register to BCBaSe 3.0 due to incomplete coverage. The Palliative Care Register captures episodes for less than half of the breast cancer patients who died and only includes the last care episodes preceding death. The unit costs for palliative care were extracted from a Swedish end-of-life study on cancer patients, without being specific to breast cancer.^19^ To avoid double-counting costs, we assumed that half of the costs were attributed to inpatient/outpatient care episodes. Secondly, pharmaceutical costs were undervalued since we did not include all hospital-based requisition drugs in the total estimate and no other drugs than those specific for breast cancer. Due to confidentiality issue and data availability, some hospital-based drugs at the brand level cannot be disclosed and indication splitting for breast cancer was not available. Additionally, DRG for outpatient care regarding intravascular drug delivery was not fully captured by BCBaSe 3.0. Costs of the hospital-based requisition drugs could be further investigated when such data are available. Thirdly, we did not include breast cancer costs within primary care due to limited data availability.

Although we have underestimated some costs, given a higher prevalence, the estimated total cost of breast cancer is higher than the total costs for other types of cancer in Sweden.^22,30,31^ The mean cost per capita in Sweden for breast cancer has risen to 51 euro, compared to the European study (10 euro, 2009 price).^9^ In Europe, breast cancer has the second highest cost among different cancer types, exceeded only by lung cancer.^9^ Our study confirms that breast cancer is a major public health concern and represents a large economic burden in Sweden. From a policy perspective, implementing more economically efficient screening strategies, along with more cost-effective treatment modalities, could contribute to greater economic sustainability by enabling earlier breast cancer detection and improved survival outcomes.

## Conclusion

Breast cancer has an annual total societal cost of 526 million and constitutes a massive economic burden in Sweden. By studying the mean cost per patient-year by subgroups, we have been able to provide cost estimates for future health economic evaluations of breast cancer related health care services and interventions.

## Data Availability

All data produced in the present work are contained in the manuscript.

## Notes

### Competing Interest Statement

The authors have declared no competing interest.

### Funding Statement

This study was funded by the Swedish Research Council, VINNOVA, Swedish Cancer Society, and the European Union.

### Author Declarations

For privacy protection, we undertook all analyses under the conditions of ethical approvals provided by the Swedish Ethical Review Authority (with approvals dnr 2019-02610 and dnr 2020-00886). Specifically, any analyses of potentially identifiable data were undertaken on secure network facilities with no reporting of results for individuals or small groups. For datasets with individual-level data, the datasets used an encrypted identifier and were pseudo-anonymised.

