## Supplementary material for "Breaking down the costs for breast cancer: Insights from Sweden’s National Quality Register"

**Appendix A. Methods**

**A1. Breast cancer subtype**

**Supplementary Table 1. Definition of breast cancer subtype**

| **Subtype** | **ER** | **PR** | **HER2** | **NHG** |
| --- | --- | --- | --- | --- |
| Luminal A like | Any  Positive | Positive  Negative | Negative  Negative | 1 or 2  1 |
| Luminal B like | Any  positive | Positive  Negative | Negative  Negative | 3  2 or 3 |
| Luminal HER2+ | Any  Positive | Positive  Negative | Positive  Positive | Any  Any |
| HER2+ | Negative | Negative | Positive | Any |
| Triple negative | Negative | Negative | Negative | Any |

ER: Estrogen receptor; PR: Progesterone receptor; HER2: Human epidermal growth factor receptor 2; NHG: Nottingham Histologic Grade

**A2. Breast cancer stage**

Breast cancer stage at diagnosis was defined using the TNM classification system 8^th^ edition combining both clinical and pathological variables in BCBaSe 3.0.^1^

**Supplementary Table 2. Definition of breast cancer stage**

| **Stage** | **T** | **N** | **M** |
| --- | --- | --- | --- |
| Stage 0 | Tis | 0 | 0 |
| Stage I | 0 or 1 | 0 | 0 |
| Stage II | 0 or 1  2  3 | 1  0 or 1  0 | 0  0  0 |
| Stage III | 0, 1, 2 or 3  3  4  Any | 2  1  0, 1 or 2  3 | 0  0  0  0 |
| Stage IV | Any | Any | 1 |

T: Tumor; N: Nodes; M: Metastasis; Tis: Carcinoma in situ

**A3. Disease state imputation**

For breast cancer patients who did not die from breast cancer, we assumed that missing M stage at diagnosis was M0. For breast cancer deaths, imputation was performed for missing M stage at diagnosis and for missing date of distant recurrence. To predict missing M stage, we used logistic regression with covariates for the log(survival time) and the age at diagnosis of breast cancer for those death cases who had complete data in M stage. We imputed five times. For each imputed dataset, we further imputed for missing dates of distant recurrence. Two flexible parametric survival models were performed using complete data: M0 to distant recurrence; and distant recurrence to death. The imputed date of recurrence was based on the probability density function (PDF) implied by the convolution of the PDFs from the two survival models using numerical quadrature.

**A4. Analysis of mean costs per patient-year**

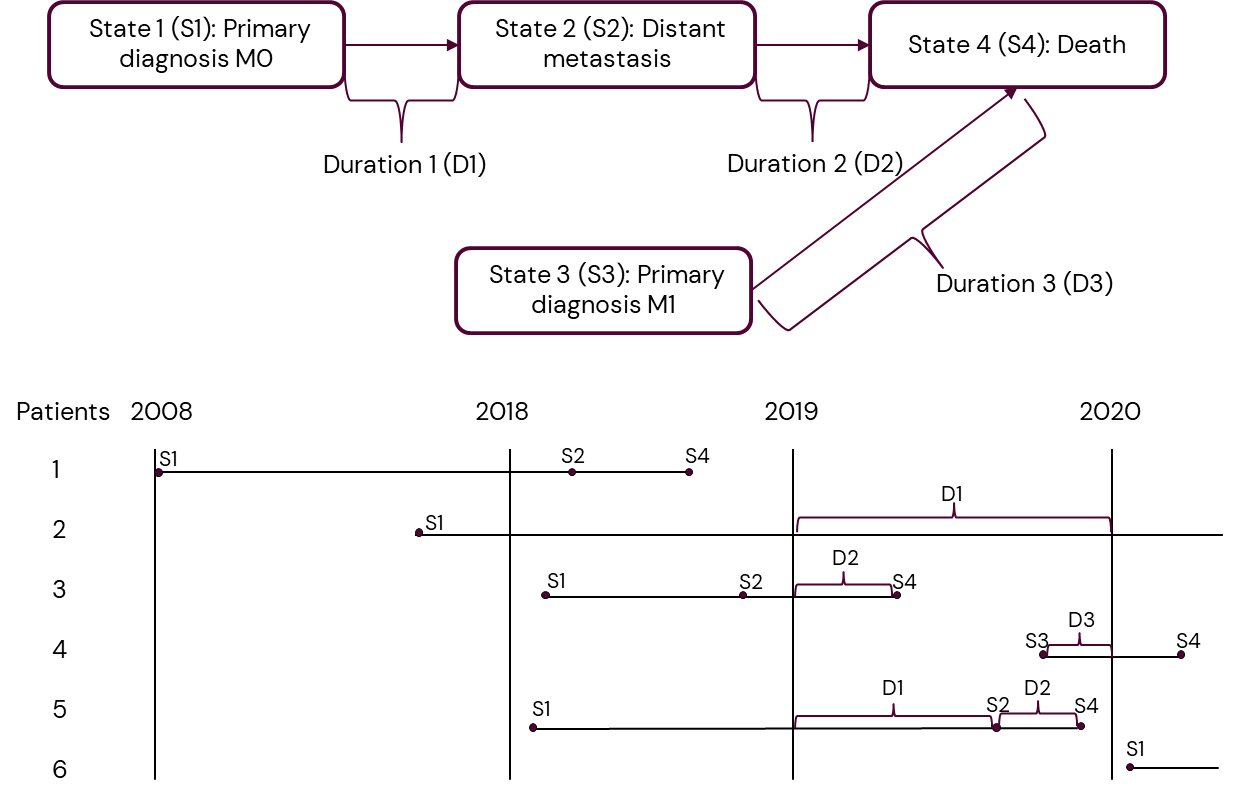

**Supplementary Figure 1. Illustration of the analysis of mean costs per patient-year, taking disease state as an example**

Patient 1 and patient 6 do not contribute to estimate.

**A5. Inpatient and outpatient care: diagnosis related group for breast cancer in Sweden**

Resource uses for inpatient and outpatient care were identified using Nordic Diagnostic Related Groups (NordDRG).

(a) For diagnosed cases, DRGs were combined with a diagnosis of invasive or non-invasive breast cancer (C50 & D05, The International Classification of Diseases, 10th version, ICD-10) and Major Diagnostic Category (MDC) of 23, 30 or 40;

**Supplementary Table 3. DRG included in the study combining a diagnosis of breast cancer and a Major Diagnostic Category in Sweden**

| **ICD** | **MDC** | **DRG - inpatient care (Swedish)** | **DRG- inpatient care (English)** |
| --- | --- | --- | --- |
| C50/D05 | 30  Breast Diseases | K01N- Mastektomi med rekonstruktion eller annan plastik | Mastectomy with reconstruction or other plastic surgery |
|  |  | K02N- Total mastektomi för malign tumör | Total mastectomy for malignant tumor |
|  |  | K03C- Rekonstruktion eller annan plastik av bröst, komplicerat | Reconstruction or other plastic surgery of breasts, complicated |
|  |  | K03E- Rekonstruktion eller annan plastik av bröst, ej komplicerat | Reconstruction or other plastic surgery of the breast, not complicated |
|  |  | K04N- Subtotal mastektomi för malign tumör | Partial mastectomy for malignant tumor |
|  |  | K10N- Bröstoperationer för benignt tillstånd utom biopsi & lokal excision | Breast surgery for benign condition except biopsy & local excision |
|  |  | K11N- Bröstbiopsi & lokal excision för benignt tillstånd | Breast biopsy & local excision for benign condition |
|  |  | K19N- Andra operationer vid bröstkörtelsjukdom | Other surgeries for mammary gland disease |
|  |  | K20C- Maligna bröstsjukdomar, komplicerat | Malignant breast disease, complicated |
|  |  | K20E- Maligna bröstsjukdomar, ej komplicerat | Malignant breast disease, not complicated |
|  |  | K30N- Benigna bröstsjukdomar | Benign breast diseases |
|  |  | K69L- Slutenvård för sjukdomar i bröstkörtel, primärvård | Inpatient care for diseases of the mammary gland, primary care |
| C50/D05 | 23  Other and unspecified health problems | W58N- Rehab av annat skäl UNS | Rehabilitation for other reasons |
| **ICD** | **MDC** | **DRG- outpatient care (Swedish)** | **DRG- outpatient care (English)** |
| C50/D05 | 30  Breast Diseases | K02O- Total mastektomi för malign tumör, öppenvård | Total mastectomy for malignant tumor, outpatient care |
|  |  | K03O- Rekonstruktion eller annan plastik av bröst, öppenvård | Reconstruction or other plastic surgery of breasts, outpatient care |
|  |  | K04O- Subtotal mastektomi för malign tumör, öppenvård | Partial mastectomy for malignant tumor, outpatient care |
|  |  | K10O- Bröstoperationer för benignt tillstånd utom biopsi & lokal excision, öppenvård | Breast surgeries for benign condition except biopsy & local excision, outpatient care |
|  |  | K11O- Bröstbiopsi & lokal excision för benignt tillstånd, öppenvård | Breast biopsy & local excision for benign condition, outpatient care |
|  |  | K19O- Andra operationer vid bröstkörtelsjukdom, öppenvård | Other operations for mammary gland disease, outpatient care |
|  |  | K20O- Läkarbesök vid maligna bröstsjukdomar | Doctor visits for malignant breast diseases |
|  |  | K20S- Läkarbesök vid maligna bröstsjukdomar, komplicerat primärvård | Doctor's visit for malignant breast diseases, complication, primary care |
|  |  | K20T- Läkarbesök vid maligna bröstsjukdomar, ej komplicerat primärvård | Doctor's visit for malignant breast diseases, no complication, primary care |
|  |  | K75O- Incision/punktion bröst, besök | Incision / puncture breast, visit |
|  |  | K89O- Läkemedelstillförsel intravasalt vid bröstkörtelsjukdomar, öppenvård | Intravascular drug delivery for mammary gland diseases, outpatient care |
|  |  | K89R- Läkemedelstillförsel intravasalt vid bröstkörtelsjukdomar, primärvård | Intravascular drug delivery for mammary gland diseases, primary care |
|  |  | K93R- Hembesök för sjukdomar i bröstkörtel, primärvård | Home visit for diseases of the mammary gland, primary care |
|  |  | K97O- Sjuksköterskebesök vid bröstkörtelsjukdomar | Nurse visits for mammary gland diseases |
|  |  | K99O- Övriga läkarbesök vid bröstkörtelsjukdomar | Other doctor visits for mammary gland diseases |
|  |  | K99R- Övriga läkarbesök vid bröstkörtelsjukdomar, primärvård | Other doctor visits for mammary gland diseases, primary care |
|  |  | K99U- Teambesök/konferens vid bröstkörtelsjukdomar, primärvård | Team visit / conference for mammary gland diseases, primary care |
|  |  | K99W- Läkarvård på distans vid bröstkörtelsjukdomar, primärvård | Remote medical care for mammary gland diseases, primary care |
|  |  | K99X- Teambesök/konferens vid bröstkörtelsjukdomar | Team visit / conference for mammary gland diseases |
|  |  | K99Z- Läkarvård på distans vid bröstkörtelsjukdomar | Remote medical care for mammary gland diseases |
| C50/D05 | 40  Overall Problems in Outpatient Care | X05O Implantation av pump el injektionsport, öppenvård | Implantation of pump or injection port, outpatient care |
|  |  | X11O Strålbehandling, resurskrävande, öppenvård | Radiotherapy, resource-intensive, outpatient care |
|  |  | X12O Strålbehandling inklusive förberedande åtgärder, öppenvård | Radiotherapy including preparatory measures, outpatient care |
|  |  | X13O Brakyterapi och isotopbehandling, öppenvård | Brachytherapy and radionuclide therapy, outpatient care |
|  |  | X14O Strålbehandling, mindre resurskrävande, öppenvård | Radiotherapy, less resource-intensive, outpatient care |
|  |  | X15O Strålbehandlingsförberedelse | Radiotherapy preparation |
|  |  | X41O Skintigrafier, besök | Scintigraphy, visit |
|  |  | X51O Venkatetrar, besök | Venous catheters, visit |

(b) To account for diagnostic procedures for patients who were clinically diagnosed, we compared the numbers of DRGs occurred within 1 month prior to a diagnosis of breast cancer to 1 year prior to the diagnosis. McNemar's test with Bonferroni correction was performed to test the differences between the numbers of patients having a DRG in each period. We selected those DRGs that had statistically significant change (after Bonferonni correction) in the numbers of patients and are relevant to a breast cancer diagnosis (Supplementary Table 4). We assumed those episodes within a month prior to the diagnosis were associated with breast cancer. DRGs occurred in the year of 2019 were included in the analysis.

**Supplementary Table 4. DRG included for diagnostic procedures prior to the diagnosis for patients who were clinically diagnosed**

| **MDC** | **DRG - inpatient care (Swedish)** | **DRG- inpatient care (English)** |
| --- | --- | --- |
| 30  Breast Diseases | K10N- Bröstoperationer för benignt tillstånd utom biopsi & lokal excision | Breast surgery for benign condition except biopsy & local excision |
| **MDC** | **DRG- outpatient care (Swedish)** | **DRG- outpatient care (English)** |
| 30  Breast Diseases | K04O- Subtotal mastektomi för malign tumör, öppenvård | Partial mastectomy for malignant tumor, outpatient care |
|  | K10O- Bröstoperationer för benignt tillstånd utom biopsi & lokal excision, öppenvård | Breast surgeries for benign condition except biopsy & local excision, outpatient care |
|  | K11O- Bröstbiopsi & lokal excision för benignt tillstånd, öppenvård | Breast biopsy & local excision for benign condition, outpatient care |
|  | K20O- Läkarbesök vid maligna bröstsjukdomar | Doctor visits for malignant breast diseases |
|  | K75O- Incision/punktion bröst, besök | Incision / puncture breast, visit |
|  | K99O- Övriga läkarbesök vid bröstkörtelsjukdomar | Other doctor visits for mammary gland diseases |
| 17  Myeloproliferative diseases and unspecified neoplasms | R99O Övriga läkarbesök vid ospecifika tumörsjukdomar och blodcancer | Other doctor visits for non-specific tumor diseases and blood cancer |

**A6. ATC code identification for pharmaceutical use**

We were able to identify 36 substances with indications for breast cancer, based on information from fass.se (Supplementary Table 3). Unit costs using sales values (Apoteksaktörens utförsäljningspris, AUP) for 190 drugs by brand, package and strength were extracted using 2023 prices and assigned in the following steps:

(a) for drugs included on the high-cost protection list (högkostnadsskydd), the historical costs in 2023 were extracted from The Dental and Pharmaceutical Benefits Agency database^2^;
(b) for drugs not included on the high-cost protection list, costs were extracted from the pharmacy websites (extracted in 2023-06)^3^;
(c) if AUP values were not available from TLV or pharmacy websites, unit costs were given the mean values based on other drugs under the same chemical substance with the same strength and package;
(d) if information on strength and package was missing, AUP values remained missing.

**Supplementary Table 5. List of drugs by chemical substance for breast cancer in Sweden in 2019**

| **ATC 5** | **Substans (Swedish)** | **Substance (English)** | **Class** | **Sub-class** | **Multiple indication** |
| --- | --- | --- | --- | --- | --- |
| L01BC02 | Fluorouracil | Fluorouracil | Chemotherapy | Antimetabolite | Yes |
| L01DB03 | Epirubicin | Epirubicin | Chemotherapy | Anthracycline | Yes |
| L01DB07 | Mitoxantron | Mitoxantrone | Chemotherapy | Anthracenedione | Yes |
| L01AA01 | Cyklofosfamid | Cyclophosphamide | Chemotherapy | Alkylating agent | Yes |
| L01CD02 | Docetaxel | Docetaxel | Chemotherapy | Taxane | Yes |
| L01CD01 | Paklitaxel | Paclitaxel | Chemotherapy | Taxane | Yes |
| L01BC06 | Kapecitabin | Capecitabine | Chemotherapy | Antimetabolite | Yes |
| L01DB01 | Doxorubicin | Doxorubicin | Chemotherapy | Anthracycline | Yes |
| L01XX41 | Erbulin | Erbulin | Chemotherapy | Microtubule inhibitor | Yes |
| L01CA04 | Vinorelbin | Vinorelbine | Chemotherapy | Vinca alkaloid | Yes |
| L01XA02 | Karboplatin | Carboplatin | Chemotherapy | Platinum compound | Yes |
| L01BA01 | Metotrexat | Methotrexate | Chemotherapy | Antimetabolite | Yes |
| L01BC05 | Gemcitabin | Gemcitabine | Chemotherapy | Antimetabolite | Yes |
| L01XK01 | Olaparib | Olaparib | Targeted therapy | PARP Inhibitor | Yes |
| L01XK04 | Talazoparib | Talazoparib | Targeted therapy | PARP inhibitor | Yes* |
| L02BG04 | Letrozol | Letrozole | Hormonal Therapy | Aromatase Inhibitor | No |
| L01EF01 | Palbociklib | Palbociclib | Targeted therapy | CDK4/6 Inhibitor | No |
| L01EF02 | Ribociklib | Ribociclib | Targeted therapy | CDK4/6 Inhibitor | No |
| L01EF03 | Abemaciklib | Abemaciclib | Targeted therapy | CDK4/6 Inhibitor | No |
| L02BA03 | Fulvestrant | Fulvestrant | Hormonal Therapy | Estrogen Receptor Antagonist | No |
| L01EG02 | Everolimus | Everolimus | Targeted therapy | mTOR Inhibitor | Yes |
| L02AB01 | Megestrolacetat | Megestrol acetate | Hormonal Therapy | Progestin | No |
| L02AB02 | Medroxiprogesteronacetat | Medroxyprogesterone acetate | Hormonal Therapy | Progestin | Yes |
| L02BG06 | Exemestan | Exemestane | Hormonal Therapy | Aromatase Inhibitor | No |
| L02BG03 | Anastrozol | Anastrozole | Hormonal Therapy | Aromatase Inhibitor | No |
| L02BA01 | Tamoxifen | Tamoxifen | Hormonal Therapy | Aromatase Inhibitor | No |
| L02BA02 | Toremifen | Toremifene | Hormonal Therapy | SERM | No |
| L01FF02 | Pembrolizumab | Pembrolizumab | Immunotherapy | Checkpoint Inhibitor | Yes |
| L02AE03 | Goserelin | Goserelin | Hormonal Therapy | LHRH Agonist | Yes |
| L02AE04 | Triptorelin | Triptorelin | Hormonal Therapy | LHRH Agonist | Yes |
| L01FD01 (L01XC03) | Trastuzumab | Trastuzumab | Targeted therapy | HER2 targeted | No** |
| L01FD02 (L01XC13) | Pertuzumab | Pertuzumab | Targeted therapy | HER2 targeted | No |
| L01FD03 | Trastuzumab emtansin | Trastuzumab-emtansine | Targeted therapy | HER2 targeted | No |
| L01EH02 | Neratinib | Neratinib | Targeted therapy | HER2 targeted | No |
| L01EH01 | Lapatinib | Lapatinib | Targeted therapy | HER2 targeted | No |
| L01FF05 | Atezolizumab | Atezolizumab | Immunotherapy | Checkpoint Inhibitor | Yes |

ATC: Anatomical Therapeutic Chemical Classification System; LHRH: Luteinizing hormone-releasing hormone; HER2: Human epidermal growth factor receptor 2; PARP: Poly (ADP‐ribose) polymerase; mTOR: mammalian target of rapamycin

*Drug under this substance was approved in 2019 for breast cancer only.

** Certain brands under this substance have multiple indications

**A7. Informal care**

Resource use for informal care was estimated using data from WAVE 8 (release version 8.0.0) of the Survey of Health, Ageing and Retirement in Europe (SHARE) project.^4^

Hours of informal care for patients severely limited in daily activities were estimated by adding the age-group products of:

(a) Breast cancer case that had not died in 2019 in Sweden, from BCBaSe 3.0;
(b) Predicted probability of being severely limited in daily activities due to cancer: logistic regression after adjusting for age, sex, presence of cancer, presence of comorbidities and country of residence, from WAVE8 of SHARE;
(c) Predicted probability of having comorbidities: logistic regression after adjusting for age, sex, presence of cancer, and country of residence, from WAVE8 of SHARE;
(d) Predicted probability of receiving informal care due to cancer: logistic regression after adjusting for age, sex, presence of cancer, presence of comorbidities and country of residence for care from inside and outside the household respectively, from WAVE8 of SHARE;
(e) Predicted number of hours of informal care received: linear regression adjusting for age, sex, presence of cancer, presence of comorbidities and country of residence for care from inside and outside the household respectively, from WAVE8 of SHARE. We assumed 1 hour/day from outside household and 1.1 hour/day from inside household based on a previous Swedish study^5^;
(f) Predicted probability of care provider at working age: linear regression adjusting for age, sex, presence of cancer, presence of comorbidities and country of residence, from WAVE8 of SHARE.

Hours of informal care for patients in terminal illness were estimated by adding the age-group products of:

(a) Number of death due to breast cancer in 2019 in Sweden, from BCBaSe 3.0;
(b) Predicted probability of obtaining informal care in the last year before death from cancer: logistic regression after adjusting for age, sex, cancer as cause of death and country of residence, from WAVE8 of SHARE;
(c) Predicted number of total hours of informal care received due to cancer: linear regression after adjusting for age, sex, cancer as cause of death and country of residence, from WAVE8 of SHARE; For those who did not report hours but provided frequency of care provided, we assumed 15 days of care for “less 1 month”, 60 days of care for “1-3 months”, 135 days of care for “3-6 months”, 270 days of care for “6-12 month”, and 360 days of care for “a full year”.
(d) Predicted probability of care provider at working age: linear regression adjusting for age, sex, cancer as main cause of death and country of residence, using data from WAVE8 of SHARE.

**Appendix B. Results:**

**B1. Study population by breast cancer subtype and age group**

**Supplementary Table 6. BCBaSe 3.0 prevalent cases distribution by breast cancer subtype and age group**

| **Age group**  **at diagnosis**  **Breast cancer**  **subtype** | **< 40** | **40-49** | **50-59** | **60-69** | ≥**70** | **Total** |
| --- | --- | --- | --- | --- | --- | --- |
| **Luminal A like** | 703 | 5,570 | 8,165 | 13,284 | 10,756 | 38,478 |
| **Luminal B like** | 456 | 1,640 | 2,564 | 4,026 | 3,643 | 12,329 |
| **Luminal HER2+** | 575 | 1,310 | 1,569 | 1,755 | 1,339 | 6,548 |
| **HER2+** | 248 | 512 | 823 | 834 | 545 | 2,962 |
| **Triple negative** | 618 | 986 | 1,291 | 1,547 | 1,358 | 5,800 |
| **Subtype missing** | 708 | 3,126 | 3,532 | 4,617 | 4,860 | 16,843 |
| **Total** | 3,308 | 13,144 | 17,944 | 26,063 | 22,501 | 82,960 |

HER2: Human epidermal growth factor receptor 2

**B2. Unit costs and quantity of resource utilisation based on BCBaSe 3.0**

**Supplementary Table 7. Unit costs and quantity of resource utilisation for breast cancer, based on BCBaSe 3.0 (2023 price, euro)**

| **Inpatient care diagnosis-related group** | **No. Patients** | **No. Episodes** | **Mean cost/patient** | |
| --- | --- | --- | --- | --- |
| K01N Mastectomy with reconstruction or other plastic surgery | 345 | 345 | 9,707 | |
| K02N Total mastectomy for malignant tumor | 1,948 | 1,952 | 6,603 | |
| K03C Reconstruction or other plastic surgery of breasts, complicated | 89 | 89 | 16,681 | |
| K03E Reconstruction or other plastic surgery of the breast, not complicated | 125 | 125 | 8,327 | |
| K04N Partial mastectomy for malignant tumor | 1,246 | 1,278 | 6,007 | |
| K10N Breast surgery for benign condition except biopsy & local excision | 1 | 1 | 5,785 | |
| K19N Other surgeries for mammary gland disease | 136 | 136 | 5,092 | |
| K20C Malignant breast disease, complicated | 530 | 712 | 10,067 | |
| K20E Malignant breast disease, not complicated | 529 | 706 | 5,743 | |
| K30N Benign breast diseases | 4 | 5 | 4,492 | |
| W58N Rehabilitation for other reasons | 182 | 185 | 5,664 | |
| **Outpatient care diagnosis-related group** | **No. Patients** | **No. Episodes** | **Mean cost/patient** | |
| K02O Total mastectomy for malignant tumor | 617 | 624 | 4,647 | |
| K03O Reconstruction or other plastic surgery of breasts | 793 | 802 | 4,854 | |
| K04O Partial mastectomy for malignant tumor | 3,520 | 3,742 | 3,962 | |
| K19O Other operations for mammary gland disease | 445 | 447 | 1,381 | |
| K20O Doctor visits for malignant breast diseases | 15,549 | 47,222 | 2,123 | |
| K75O Incision / puncture breast, visit | 393 | 455 | 642 | |
| K89O Intravascular drug delivery for mammary gland diseases | 1 | 1 | 825 | |
| K99O Other doctor visits for mammary gland diseases | 4,738 | 5,564 | 608 | |
| K99X Team visit / conference for mammary gland diseases | 321 | 457 | 522 | |
| X05O Implantation of pump or injection port | 371 | 382 | 1,716 | |
| X11O Radiotherapy, resource-intensive | 76 | 444 | 1,827 | |
| X12O Radiotherapy including preparatory measures | 1,753 | 7,428 | 1,437 | |
| X13O Brachytherapy and radionuclide therapy | 2 | 2 | 1,183 | |
| X14O Radiotherapy, less resource-intensive | 2,124 | 21,707 | 3,089 | |
| X15O Radiotherapy preparation | 2,481 | 2,561 | 524 | |
| X41O Scintigraphy, visit | 655 | 722 | 808 | |
| X51O Venous catheters, visit | 328 | 366 | 695 | |
| **Prescribed drugs** | **No. Patients** | **Mean cost/patient** | | |
| L01AA01 Cyclophosphamide | 84 | 32 | | |
| L01BC06 Capecitabine | 466 | 253 | | |
| L01CA04 Vinorelbine | 34 | 928 | | |
| L01CD01 Paclitaxel | 13 | 111 | | |
| L01CD02 Docetaxel | 1 | 36 | | |
| L01DB03 Epirubicin | 14 | 33 | | |
| L01DB07 Mitoxantrone | 1 | 2,948 | | |
| L01EF01 Palbociclib | 475 | 12,046 | | |
| L01EF02 Ribociclib | 187 | 3,220 | | |
| L01EF03 Abemaciclib | 13 | 2,336 | | |
| L01EG02 Everolimus | 23 | 3,092 | | |
| L01EH01 Lapatinib | 25 | 7,067 | | |
| L01XC03 Trastuzumab | 11 | 2,093 | | |
| L01XC13 Pertuzumab | 3 | 9,592 | | |
| L01XK01 Olaparib | 36 | 12,075 | | |
| L01XX41 Erbulin | 1 | 4,254 | | |
| L02AB01 Megestrol acetate | 9 | 140 | | |
| L02AB02 Medroxyprogesterone acetate | 5 | 250 | | |
| L02AE03 Goserelin | 1,198 | 1,064 | | |
| L02AE04 Triptorelin | 5 | 456 | | |
| L02BA01 Tamoxifen | 16,099 | 65 | | |
| L02BA02 Toremifene | 42 | 217 | | |
| L02BA03 Fulvestrant | 412 | 1,230 | | |
| L02BG03 Anastrozole | 7,624 | 142 | | |
| L02BG04 Letrozole | 11,999 | 190 | | |
| L02BG06 Exemestane | 1,937 | 201 | | |
| **Informal care** | **No. Hours** | | | **Mean cost/Hour** |
| Severely limited in daily activity | 1,044,321 | | | 28.5 |
| Terminal illness | 157,502 | | | 28.5 |
| **Productivity losses - morbidity** | **No. Patients** | **No. Days** | | **Mean salary/Day** |
| Sick absence (including 14-day short sick leave) | 5,690 | 832,055 | | 228 |
| Disability pension | 145 | 35,542.5 | | 228 |
| **Productivity losses - mortality** | **No. Patients** | **No. Years** | | **Gross earning/Year** |
| Pre-mature mortality | 297 | 280.18 | | 57,240 |

**B3. Prescribed drugs by drug class**

**Supplementary Table 8. Cost of prescribed drugs by drug class, based on BCBaSe 3.0 (2023 price, euro)**

| **Drug class** | **Total cost (€)** | **No. Patients** |
| --- | --- | --- |
| **Chemotherapy** | **161,333** | **541** |
| Alkylating agent | 2,691 | 84 |
| Anthracenedione | 2,948 | 1 |
| Anthracycline | 462 | 14 |
| Antimetabolite | 117,936 | 466 |
| Microtubule inhibitor | 4,254 | 1 |
| Taxane | 1,479 | 14 |
| Vinca alkaloid | 31,564 | 34 |
| **Hormone therapy** | **6,595,952** | **35,802** |
| Aromatase Inhibitor | 4,801,042 | 35,433 |
| Estrogen Receptor Antagonist | 506,919 | 412 |
| LHRH Agonist | 1,276,352 | 1,203 |
| Progestin | 2,516 | 13 |
| Selective Estrogen Receptor Modulator | 9,123 | 42 |
| **Targeted therapy** | **7,088,457** | **745** |
| CDK4/6 Inhibitor | 6,354,168 | 657 |
| HER2 targeted | 228,471 | 36 |
| PARP Inhibitor | 434,700 | 36 |
| mTOR Inhibitor | 71,118 | 23 |

LHRH: Luteinizing hormone-releasing hormone; HER2: Human epidermal growth factor receptor 2; PARP: Poly (ADP‐ribose) polymerase; mTOR: mammalian target of rapamycin

**B4. Breakdown of direct costs by subgroup**

**Supplementary Table 9. Costs of inpatient/outpatient care and prescribed drugs per patient-year, by subgroups (2023 price, euro)**

| **Type of care** | **Subgroup** | **First year of follow-up** | |  | **Second and subsequent years of follow-up** | | | **Total years of follow up** | | | | | | |
| --- | --- | --- | --- | --- | --- | --- | --- | --- | --- | --- | --- | --- | --- | --- |
|  |  | **Total cost (€)** | **Total patient-years** | **Cost/patient-year (€)** | **Total cost (€)** | **Total patient-years** | **Cost/patient-year (€)** | **Total cost (€)** | | **Total patient-years** | | | | **Cost/patient-year (€)** |
| **Inpatient care** | **Breast cancer subtype** | | |  |  |  |  | |  | |  |  |  | |
|  | Luminal A like | 9,828,335 | 3,462 | 2,839 | 1,642,308 | 32,331 | 51 | 11,470,643 | | 35,793 | | | | 320 |
|  | Luminal B like | 4,620,931 | 1,200 | 3,851 | 811,434 | 10,202 | 80 | 5,432,365 | | 11,402 | | | | 476 |
|  | Luminal HER2+ | 2,817,811 | 680 | 4,144 | 596,405 | 5,384 | 111 | 3,414,216 | | 6,064 | | | | 563 |
|  | HER2+ | 1,500,502 | 301 | 4,987 | 267,906 | 2,419 | 111 | 1,768,408 | | 2,720 | | | | 650 |
|  | Triple negative | 2,792,150 | 671 | 4,160 | 496,560 | 4,559 | 109 | 3,288,710 | | 5,230 | | | | 629 |
|  | **Breast cancer stage** | | | | | | | | | | | | | |
|  | Stage 0 | 1,222,788 | 561 | 2,180 | 293,781 | 5,014 | 59 | 1,516,569 | | 5,575 | | | | 272 |
|  | Stage I | 7,487,218 | 3,506 | 2,136 | 2,092,126 | 33,365 | 63 | 9,579,344 | | 36,870 | | | | 260 |
|  | Stage II | 13,712,035 | 3,255 | 4,213 | 3,019,450 | 24,217 | 125 | 16,731,485 | | 27,472 | | | | 609 |
|  | Stage III | 4,303,816 | 670 | 6,422 | 1,590,541 | 5,449 | 292 | 5,894,357 | | 6,119 | | | | 963 |
|  | Stage IV | 1,006,386 | 191 | 5,262 | 762,026 | 563 | 1,354 | 1,768,412 | | 754 | | | | 2,345 |
|  | **Age group** |  |  |  |  |  |  | |  | |  |  |  | |
|  | <40 | 1,611,459 | 283 | 5,685 | 285,709 | 999 | 286 | 1,897,168 | | 1,282 | | | | 1,480 |
|  | 40-49 | 4,540,623 | 1,102 | 4,121 | 1,017,419 | 5,342 | 190 | 5,558,042 | | 6,444 | | | | 862 |
|  | 50-59 | 5,412,322 | 1,603 | 3,376 | 1,726,127 | 13,193 | 131 | 7,138,449 | | 14,796 | | | | 482 |
|  | 60-69 | 5,933,088 | 2,125 | 2,792 | 1,756,088 | 17,390 | 101 | 7,689,176 | | 19,515 | | | | 394 |
|  | ≥70 | 10,687,776 | 3,231 | 3,258 | 3,055,839 | 32,494 | 94 | 13,743,615 | | 35,725 | | | | 385 |
|  | **Disease state** |  |  |  |  |  |  | |  | |  |  |  | |
|  | Primary diagnosis M0 | 26,944,251 | 8,122 | 3,318 | 4,604,040 | 67,802 | 68 | 31,548,292 | | 75,924 | | | | 416 |
|  | De novo metastatic M1 | 1,006,386 | 191 | 5,262 | 762,027 | 563 | 1,354 | 1,768,412 | | 754 | | | | 2,345 |
|  | Distant recurrence | 1,549,101 | 363 | 4,266 | 1,160,644 | 721 | 1,609 | 2,709,745 | | 1,085 | | | | 2,498 |
| **Outpatient care** | **Breast cancer subtype** | | | | | | |  | |  | | | |  |
|  | Luminal A like | 22,960,913 | 3,462 | 6,633 | 2,904,131 | 32,331 | 90 | 25,865,044 | | 35,793 | | | | 723 |
|  | Luminal B like | 8,708,431 | 1,200 | 7,257 | 1,163,822 | 10,202 | 114 | 9,872,253 | | 11,402 | | | | 866 |
|  | Luminal HER2+ | 6,274,739 | 680 | 9,228 | 1,116,678 | 5,384 | 207 | 7,391,417 | | 6,064 | | | | 1,219 |
|  | HER2+ | 2,752,651 | 301 | 9,148 | 508,172 | 2,419 | 210 | 3,260,823 | | 2,720 | | | | 1,199 |
|  | Triple negative | 5,882,738 | 671 | 8,764 | 921,552 | 4,559 | 202 | 6,804,290 | | 5,230 | | | | 1301 |
|  | **Breast cancer stage** | | | | | | | | | | | | | |
|  | Stage 0 | 3,050,310 | 561 | 5,437 | 579,176 | 5,014 | 116 | 3,629,486 | | 5,575 | | | | 651 |
|  | Stage I | 23,999,691 | 3,506 | 6,846 | 2,970,581 | 33,365 | 89 | 26,970,272 | | 36,870 | | | | 732 |
|  | Stage II | 24,201,152 | 3,255 | 7,436 | 3,847,902 | 24,217 | 159 | 28,049,054 | | 27,472 | | | | 1,021 |
|  | Stage III | 5,474,850 | 670 | 8,170 | 1,846,152 | 5,449 | 339 | 7,321,002 | | 6,119 | | | | 1,197 |
|  | Stage IV | 1,059,867 | 191 | 5,541 | 1,154,320 | 563 | 2,051 | 2,214,187 | | 754 | | | | 2,937 |
|  | **Age group** |  |  |  |  |  |  | |  | |  |  |  | |
|  | <40 | 2,969,182 | 283 | 10,475 | 477,545 | 999 | 478 | 3,446,727 | | 1,282 | | | | 2,688 |
|  | 40-49 | 10,019,697 | 1,102 | 9,094 | 1,282,453 | 5,342 | 240 | 11,302,150 | | 6,444 | | | | 1,754 |
|  | 50-59 | 127,49,472 | 1,603 | 7,953 | 2,176,330 | 13,193 | 165 | 14,925,802 | | 14,796 | | | | 1,009 |
|  | 60-69 | 15,816,612 | 2,125 | 7,442 | 2,293,058 | 17,390 | 132 | 18,109,670 | | 19,515 | | | | 928 |
|  | ≥70 | 17,227,769 | 3,231 | 5,332 | 4,305,307 | 32,494 | 132 | 21,533,076 | | 35,725 | | | | 603 |
|  | **Disease state** |  |  |  |  |  |  | |  | |  |  |  | |
|  | Primary diagnosis M0 | 57,557,040 | 8,122 | 7,061 | 7,595,678 | 67,802 | 112 | 65,152,718 | | 75,924 | | | | 858 |
|  | De novo metastatic M1 | 1,059,867 | 191 | 5,541 | 1,154,319 | 563 | 2,051 | 2,214,187 | | 754 | | | | 2,936 |
|  | Distant recurrence | 948,928 | 363 | 2,613 | 1,001,597 | 721 | 1,388 | 1,950,523 | | 1,085 | | | | 1,798 |
| **Prescribed drugs** | **Breast cancer subtype** | | | | | | |  | |  | | | |  |
|  | Luminal A like | 585,988 | 3,462 | 169 | 4,144,404 | 32,331 | 128 | 4,730,392 | | 35,793 | | | | 132 |
|  | Luminal B like | 230,249 | 1,200 | 192 | 2,205,987 | 10,202 | 216 | 2,436,236 | | 11,402 | | | | 214 |
|  | Luminal HER2+ | 166,217 | 680 | 244 | 1,113,093 | 5,384 | 207 | 1,279,310 | | 6,064 | | | | 211 |
|  | HER2+ | 13,602 | 301 | 45 | 111,170 | 2,419 | 46 | 124,772 | | 2,720 | | | | 46 |
|  | Triple negative | 46,093 | 671 | 69 | 123,048 | 4,559 | 27 | 169,141 | | 5,230 | | | | 32 |
|  | **Breast cancer stage** | | | | | | | | | | | | | |
|  | Stage 0 | 4,170 | 561 | 7 | 105,582 | 5,014 | 21 | 109,752 | | 5,575 | | | | 20 |
|  | Stage I | 480,993 | 3,506 | 137 | 2,970,311 | 33,365 | 89 | 3,451,304 | | 36,870 | | | | 94 |
|  | Stage II | 610,787 | 3,255 | 188 | 4,588,994 | 24,217 | 190 | 5,199,781 | | 27,472 | | | | 189 |
|  | Stage III | 4,588,994 | 670 | 382 | 2,286,136 | 5,449 | 420 | 2,542,029 | | 6,119 | | | | 416 |
|  | Stage IV | 538,801 | 191 | 2,817 | 1,504,103 | 563 | 2,673 | 2,042,904 | | 754 | | | | 2,709 |
|  | **Age group** |  |  |  |  |  |  | |  | |  |  |  | |
|  | <40 | 112,179 | 283 | 396 | 482,914 | 999 | 484 | 595,093 | | 1,282 | | | | 464 |
|  | 40-49 | 247,980 | 1,102 | 225 | 1,494,924 | 5,342 | 280 | 1,742,904 | | 6,444 | | | | 271 |
|  | 50-59 | 363,148 | 1,603 | 227 | 2,289,036 | 13,193 | 174 | 2,652,184 | | 14,796 | | | | 179 |
|  | 60-69 | 480,816 | 2,125 | 226 | 2,914,020 | 17,390 | 168 | 3,394,836 | | 19,515 | | | | 174 |
|  | ≥70 | 699,445 | 3,231 | 216 | 4,330,565 | 32,494 | 133 | 5,030,010 | | 35,725 | | | | 141 |
|  | **Disease state** |  |  |  |  |  |  | |  | |  |  |  | |
|  | Primary diagnosis M0 | 1,305,218 | 8,122 | 161 | 7,050,875 | 67,802 | 104 | 8,356,093 | | 75,924 | | | | 110 |
|  | De novo metastatic M1 | 538,801 | 191 | 2,817 | 1,504,103 | 563 | 2,673 | 2,042,904 | | 754 | | | | 2,709 |
|  | Distant recurrence | 1,068,009 | 363 | 2,941 | 1,948,021 | 721 | 2,700 | 3,016,030 | | 1,085 | | | | 2,781 |

HER2: Human epidermal growth factor receptor 2

**B5. Sensitivity analysis**

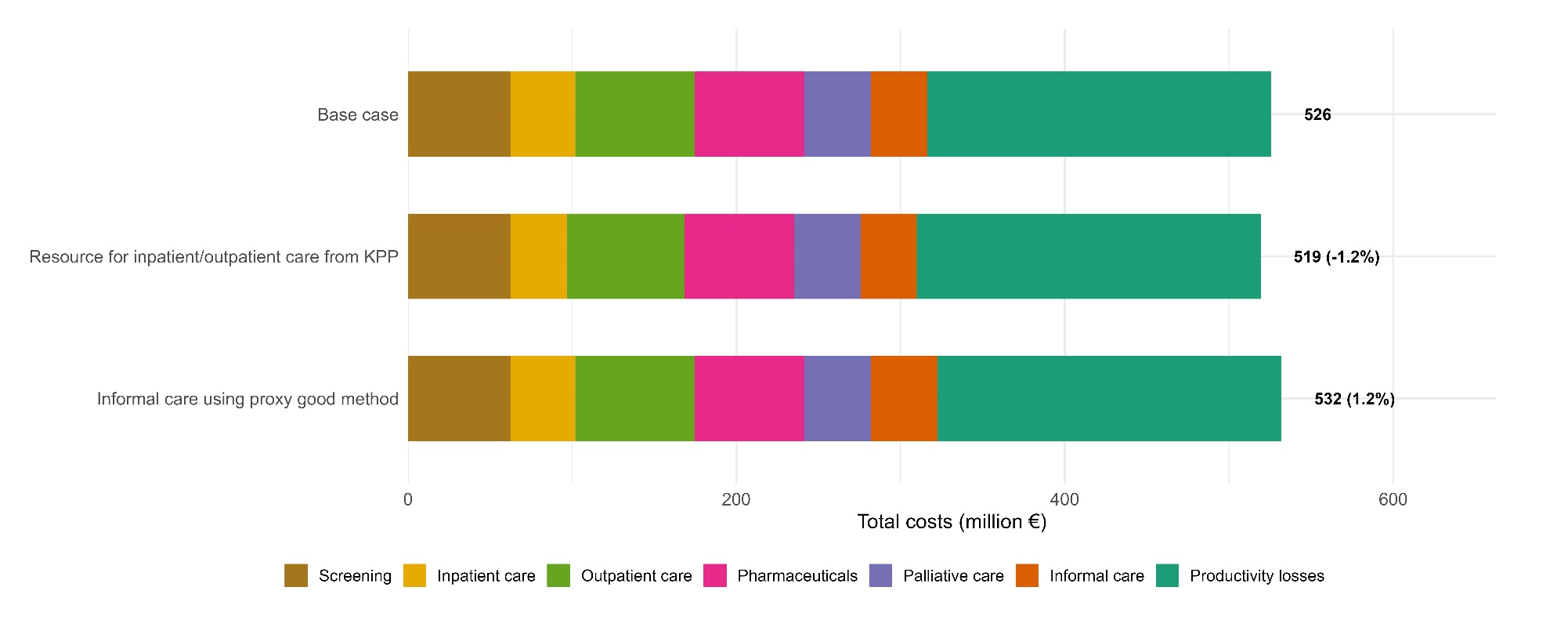

**Supplementary Figure 2. Sensitivity analyses of costs of breast cancer in Sweden**

The percentage indicates the change on the total costs compared to base case.
